# Profiles of inpatient psychiatry referrals: A 4-year analysis in a Consultation-Liaison Psychiatry Service

**DOI:** 10.1101/2025.06.26.25330384

**Authors:** Jeff Huarcaya-Victoria, David Villarreal-Zegarra, Renato D. Alarcón-Guzmán

## Abstract

**Background:** Consultation-Liaison Psychiatry (CLP) services are essential for addressing the psychiatric needs of patients with complex medical conditions in general hospitals.

**Aims:** The study aims to characterize profiles of inpatient psychiatry referrals and assess trends and correlations between referrals, patient demographics, and psychiatric and somatic diagnoses.

**Method:** Data from January 2020 to December 2023 were collected from a CLP service during and after the pandemic, using psychiatric diagnoses from the International Classification of Diseases, Tenth Revision (ICD-10). Statistical analyses, including interrupted time series analysis with four periods, and including linear and segmented analyses, plus selective use of autocorrelation tests, were conducted to examine referral patterns and their associations with socio-demographic factors.

**Results:** 6,105 patients were referred to the CLP Service during the study period, which was 6.73% of all hospital admissions. Medicine and Pneumology exhibited consistently high referrals, while services like Rheumatology and Endocrinology had lower rates. Common somatic diagnoses included neoplasms (20.7%) and respiratory diseases (9.4%), while neurotic, stress-related, and somatoform disorders were prevalent psychiatric diagnoses (42.5%). Interrupted time series analysis revealed fluctuations in monthly care visits, with notable decreases coinciding with the onset of the COVID-19 pandemic.

**Conclusions:** Our study elucidated the characteristics of patients receiving CLP services at a major Peruvian general hospital, revealing depression as a prevalent reason for consultation, and highlighting the dynamic nature of psychiatric care delivery, especially amidst the COVID-19 pandemic.

## Background

In the global context of medical practice, Consultation-Liaison Psychiatry (CLP) has evolved as a specialized field, addressing the multiple intersections of mental and physical health (1, 2). Originating in the early 20th century, the development of CLP was significantly influenced by the need to address psychiatric issues in medically ill patients, particularly those hospitalized with complex medical conditions (3, 4). Throughout the decades it has evolved to become a critical component of an integral, comprehensive healthcare that emphasizes the importance of a truly multidisciplinary approach (5, 6). There is also evidence that some CLP services have demonstrated their efficacy in various healthcare settings, contributing not only to improved patient outcomes, but also proving to be cost-effective by reducing hospital stays when implemented early in the treatment process (7). The effective integration of psychiatric consultation within general medical care has proven to be a crucial component in resource-constrained environments, where the challenges imposed by strong limitations and diverse cultural contexts demand innovative approaches.

In low and middle-income countries (LMIC) like Peru, CLP assumes even greater significance as healthcare systems face a great variety of challenges, from deficits in financial and human resources to socio-cultural barriers impacting the delivery of mental health services (8, 9). Inadequate attention to medical and psychiatric comorbidities can result in a substantial burden on individual and collective healthcare systems. Therefore, CLP research in low and middle-resource settings is not only timely but imperative to inform intervention strategies and enhance the quality of care provided to populations grappling with multiple health challenges.

The history of CLP in Peru finds its roots in the pioneering efforts of the “Grupo del Obrero”, spearheaded by Dr. Carlos Alberto Seguin early in the second half of the last Century. Seguin, who trained under the mentorship of Flanders Dunbar—a significant figure in the development of psychosomatic medicine—founded the Psychiatry Service of the then-called Labor Hospital (Hospital Obrero, in Spanish) that opened its doors in 1941 (10), and later changed its name to National Hospital Guillermo Almenara Irigoyen (HNGAI), one of the clinical establishments of the Social Security system. These connections profoundly influenced the evolution of CLP and psychosomatic medicine in Peru. Under Seguin’s leadership, the HNGAI Psychiatry Service nurtured a fruitful collaboration between psychiatrists and non-psychiatric medical professionals, between psychiatry and other fields of medicine (11). The main instrument for this achievement was, not surprisingly, an active exchange of interdisciplinary consultations and clinical presentations. As mentioned, throughout the second half of the 20^th^ Century, the Psychiatry Department at HNGAI became a trailblazer in the field of CLP in Peru, grounded on a psychosomatic medicine initially nourished by psychoanalytic foundations (12).

In the current Peruvian context, CLP adheres to regulations and guidelines established by the Ministry of Health (MINSA). Chapter V of Peru’s Mental Health Law, enacted on March 5, 2020, makes clear that mental health services in general hospitals should be organized in the form of interdisciplinary teams providing liaison psychiatry care. It also emphasizes that any mental health approach must be primarily grounded on the protection of and respect for human rights, and focused on recovery (13).

The teaching values and advantages of this multidisciplinary, integrated work in Peru have been duly recognized. Nevertheless, in spite of the pressing needs and the significant relevance that this area of mental health has reached and holds for general hospital patient populations, research productivity has been limited (14–17). The lack of resources and specific research data on CLP care poses a considerable challenge, hindering the ability to document the provision of specialized and effective care. Therefore, a comprehensive understanding of patient characteristics and the care they receive becomes an indispensable basis to enhance the quality of services and develop interventions that could improve and optimize the treatment of patients with medical conditions exhibiting concurrent mental symptoms.

This study aimed to examine the characteristics of a large sample of patients who attended over a four-year period (2020–2023) by a CLP service in a low– and middle-income country, such as Peru. The primary objectives were to describe the sociodemographic and clinical characteristics of patients assessed by the CLP service using annual and aggregated frequency and percentage analyses, to analyze the structure and characteristics of the service’s database, and to evaluate potential changes in clinical practice over time, particularly in response to external influences such as the COVID-19 pandemic. Given its descriptive nature, this objective does not involve an inferential hypothesis. Additionally, as a secondary objective, an interrupted time series analysis was conducted to assess monthly variations in psychiatric consultations using four predefined interruption periods (start of the pandemic in March 2020, January 2021, January 2022, and January 2023). We hypothesized that the COVID-19 pandemic negatively impacted the number of patient consultations conducted by the CLP service.

By including an interrupted time series analysis, we quantitatively evaluate how external disruptions influenced psychiatric service demand, offering a methodological approach not commonly applied in similar CLP studies. Additionally, our study contributes to the limited international literature on CLP services in Latin America, presenting a unique opportunity to compare psychiatric referral patterns and service utilization with those in higher-income settings. Findings from this study may have broader implications for optimizing CLP models in resource-limited environments, informing mental health policies and service delivery strategies.

## Methods

### Study design

This study employed a retrospective observational design to analyze data collected from the CLP Service of the National Hospital Guillermo Almenara Irigoyen (HNGAI), a general hospital in Lima, Peru.

### Participants

The participants were patients from the CLP Service of the HNGAI in Lima, Peru. The service provides consultation to inpatients admitted to medical or surgical services and who, concurrently present psychiatric disorders or require a thorough psychiatric assessment, diagnosis and eventual management intervention. The study timeframe spanned from January 2020 to December 2023. Utilizing a non-probabilistic sampling method, the inclusion criteria stated participants to be of legal age (>18 years), and individuals with incomplete data were to be excluded from the analyzed sample.

### Setting

As mentioned above, the HNGAI was the study site, a highly complex hospital with 960 beds, located in Lima-Peru, one of the three largest establishments administered by the Social Security system, and also a tertiary referral center for all medical specialities, including psychiatry (http://www.essalud.gob.pe/estadistica-institucional/). It provides health care services to 1,547,840 individuals with Social Security insurance. As it attends to virtually all kinds of pathological conditions, it was classified in 2015 as a Specialized Health Institute III-2, the highest level conferred by Peru’s Ministry of Health.

The Consultation-Liaison Psychiatry (CLP) Service at HNGAI responds to psychiatric consultation requests from different medical and surgical departments (17). All patient evaluations conducted by the CLP Service have been initially documented in the hospital’s electronic medical record system. Additionally, since September 2020, all patient evaluations conducted by the CLP team have been systematically recorded in a Google Form at the time of assessment. This electronic registry ensures standardized, real-time data collection, reducing reliance on retrospective chart review. Attending psychiatrists and psychiatry residents directly input clinical information into the form, ensuring data accuracy and consistency.

### Instruments and variables

#### Psychiatric diagnoses

Psychiatric diagnoses were determined by attending psychiatrists specializing in consultation-liaison psychiatry, based on clinical evaluation and documented according to the International Classification of Diseases, Tenth Revision (ICD-10) criteria (18). All psychiatric diagnoses identified during the assessment were recorded in the electronic medical record. However, only the primary diagnosis—the condition most relevant to the patient’s clinical presentation and treatment—was considered for analytical purposes.

#### Somatic diagnoses

The somatic diagnoses recorded in this study cover a wide range of medical conditions evaluated by attending physicians in the medical or surgical wards. They include but are not limited to cardiovascular diseases, respiratory conditions, gastrointestinal disorders, endocrine abnormalities, neurological entities, and infectious diseases

#### Sociodemographic covariates

Several socio-demographic variables were recorded: Age (subgroups of 18-34, 35-49, 50-64, and 65 years or older), gender (men and women), marital status (single, common law or co-habitant, married, widowed or separated), level of education (none, primary, high school, technical and university levels), current employment status (not working, working or retired), living condition (alone or with others), reason for consultation, service that requested it, recommendations of the liaison psychiatry team, and prescribed medications (antipsychotics, antidepressants, mood stabilizers, anxiolytics, etc.).

### Statistical analysis

#### Sociodemographic Characteristics

An analysis of frequencies and percentages of sociodemographic and clinical characteristics of participants by year of assessment and pooled analysis was performed.

#### Interrupted time series analysis

We used an interrupted time series analysis with four periods (pandemic onset in March 2020 [lockdown], January 2021, January 2022, and January 2023) to clarify the pandemic’s impact on the number of monthly visits during the year. Linear regression models were used to assess whether the number of monthly visits changed during the different periods. We used segmented regression analysis with Newey-West standard errors to model the data (19). Monthly time services were used. Only values with a 95% hypothesis test (p<0.05) were considered significant. Cumby-Huizinga (Breusch-Godfrey) tests were used with the ‘actest’ command to test for autocorrelation (20). Time series analyses were performed using STATA 18, and plots were generated using STATA 18 and the ggplot2 package from R Studio.

### Ethics

The Hospital Nacional Guillermo Almenara Irigoyen’s Institutional Review Board approved the protocol of this study (Carta N°122 CIEI-OIyD-GRPA-Essalud-2025). Throughout the process, the researchers had no access to identifying information about the participants. All participants were registered users of the hospital’s Liaison Psychiatry Service and received psychological or psychiatric care as needed. Participants did not need to provide informed consent, as the data was collected retrospectively. In addition, it was ensured that all data had been completely anonymized.

## Results

### Participants

During the study period, a total of 90,680 patients aged 18 years and older were admitted to the hospital. Psychiatric consultation was requested for 6,105 of them. On average, 127.2 consultations were performed per month (SD = 36.6), and the number of consultations per month is shown in Supplementary material 2. Figure 1 shows the proportion of referrals made to the liaison psychiatry service relative to the total number of referrals made in the hospital from January 2020 to December 2023. The rate started at 5.03 % in January 2020, decreased to its lowest point at 1.90% in April 2020, and then generally increased, reaching a peak of 10.50 % in November 2021. Subsequently, the rate exhibited several fluctuations, eventually declining to 5.13 % by December 2023 (see Supplementary material 2).

**Figure 1.** Proportion of referrals by month, between 2020 and 2023 (n = 6,105). Note: The denominator is the total number of referrals made in the hospital each month.

The most frequent age group was 50-64 (n=1,745; 28.6%), female (n=3,502; 57.4%), with couples (n=2,935; 58.2%), university educated (n=1,557; 32.2%), currently working (n=2,371; 49.0%), and living with someone else (n=4,598; 93.0%).

The main reasons for consultation were depression (n=1,381; 24.8%), mental state evaluation, which involves a general assessment of mental health using psychometric tests and a clinical interview (n=1,367; 24.5%), and anxiety (n=1,039; 18.6%). The main service requesting consultations was Medicine (n=1,639; 26.8%), the most frequent Liaison Psychiatry team recommendation was initiation of medication (n=3,473; 62.3%), and the most frequently prescribed medications were anxiolytics (n=2,548; 41.7%) and antidepressants (n=2,176; 35.6%). Sociodemographic characteristics are shown in Table 1.

**Table 1.**
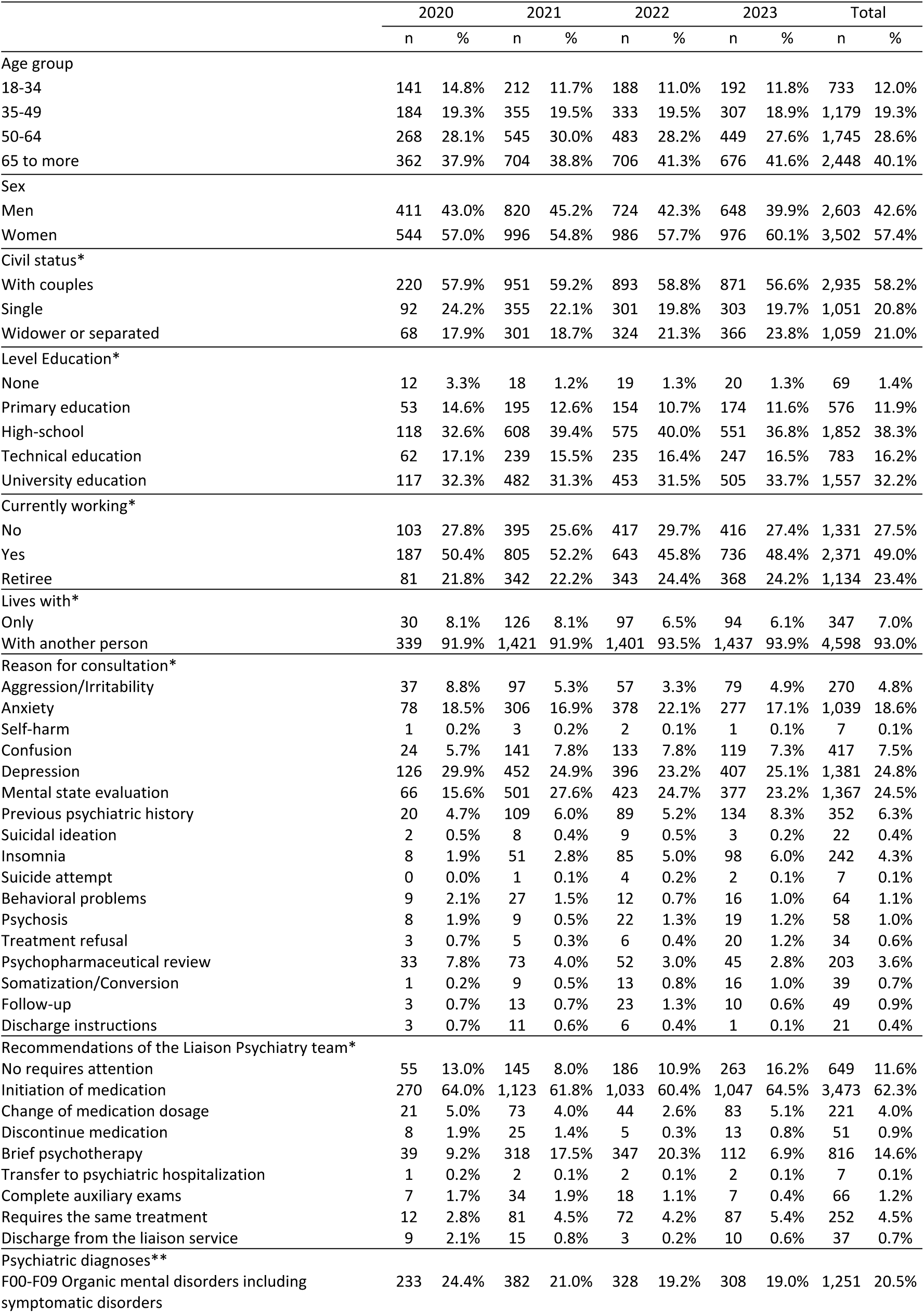
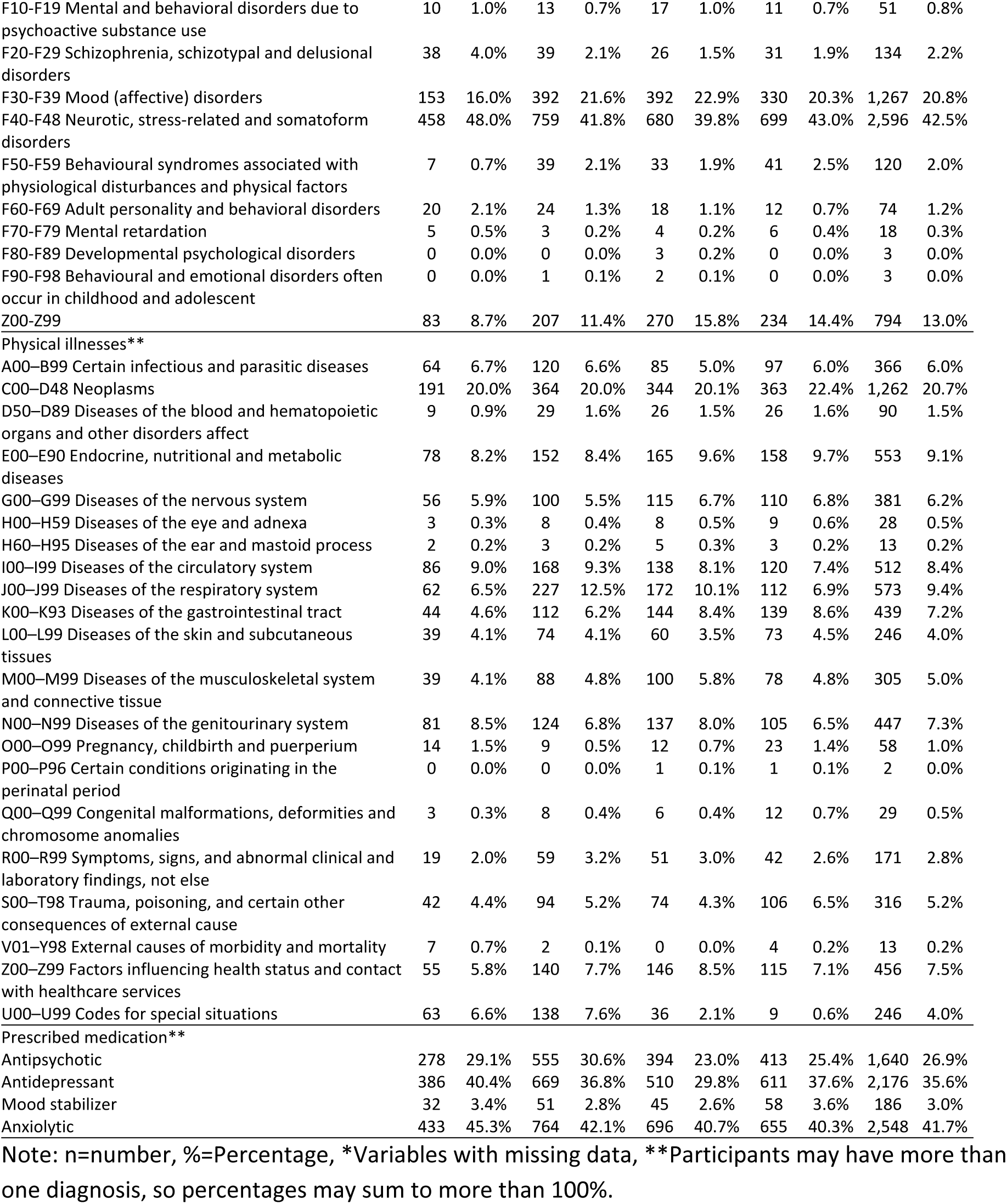
Sociodemographic characteristics of participants (n=6,105).

|  | 2020 |  | 2021 |  | 2022 |  | 2023 |  | Total |  |
| --- | --- | --- | --- | --- | --- | --- | --- | --- | --- | --- |
|  | n | % | n | % | n | % | n | % | n | % |
| Age group |  |  |  |  |  |  |  |  |  |  |
| 18-34 | 141 | 14.8% | 212 | 11.7% | 188 | 11.0% | 192 | 11.8% | 733 | 12.0% |
| 35-49 | 184 | 19.3% | 355 | 19.5% | 333 | 19.5% | 307 | 18.9% | 1,179 | 19.3% |
| 50-64 | 268 | 28.1% | 545 | 30.0% | 483 | 28.2% | 449 | 27.6% | 1,745 | 28.6% |
| 65 to more | 362 | 37.9% | 704 | 38.8% | 706 | 41.3% | 676 | 41.6% | 2,448 | 40.1% |
| Sex |  |  |  |  |  |  |  |  |  |  |
| Men | 411 | 43.0% | 820 | 45.2% | 724 | 42.3% | 648 | 39.9% | 2,603 | 42.6% |
| Women | 544 | 57.0% | 996 | 54.8% | 986 | 57.7% | 976 | 60.1% | 3,502 | 57.4% |
| Civil status* |  |  |  |  |  |  |  |  |  |  |
| With couples | 220 | 57.9% | 951 | 59.2% | 893 | 58.8% | 871 | 56.6% | 2,935 | 58.2% |
| Single | 92 | 24.2% | 355 | 22.1% | 301 | 19.8% | 303 | 19.7% | 1,051 | 20.8% |
| Widower or separated | 68 | 17.9% | 301 | 18.7% | 324 | 21.3% | 366 | 23.8% | 1,059 | 21.0% |
| Level Education* |  |  |  |  |  |  |  |  |  |  |
| None | 12 | 3.3% | 18 | 1.2% | 19 | 1.3% | 20 | 1.3% | 69 | 1.4% |
| Primary education | 53 | 14.6% | 195 | 12.6% | 154 | 10.7% | 174 | 11.6% | 576 | 11.9% |
| High-school | 118 | 32.6% | 608 | 39.4% | 575 | 40.0% | 551 | 36.8% | 1,852 | 38.3% |
| Technical education | 62 | 17.1% | 239 | 15.5% | 235 | 16.4% | 247 | 16.5% | 783 | 16.2% |
| University education | 117 | 32.3% | 482 | 31.3% | 453 | 31.5% | 505 | 33.7% | 1,557 | 32.2% |
| Currently working* |  |  |  |  |  |  |  |  |  |  |
| No | 103 | 27.8% | 395 | 25.6% | 417 | 29.7% | 416 | 27.4% | 1,331 | 27.5% |
| Yes | 187 | 50.4% | 805 | 52.2% | 643 | 45.8% | 736 | 48.4% | 2,371 | 49.0% |
| Retiree | 81 | 21.8% | 342 | 22.2% | 343 | 24.4% | 368 | 24.2% | 1,134 | 23.4% |
| Lives with* |  |  |  |  |  |  |  |  |  |  |
| Only | 30 | 8.1% | 126 | 8.1% | 97 | 6.5% | 94 | 6.1% | 347 | 7.0% |
| With another person | 339 | 91.9% | 1,421 | 91.9% | 1,401 | 93.5% | 1,437 | 93.9% | 4,598 | 93.0% |
| Reason for consultation* |  |  |  |  |  |  |  |  |  |  |
| Aggression/Irritability | 37 | 8.8% | 97 | 5.3% | 57 | 3.3% | 79 | 4.9% | 270 | 4.8% |
| Anxiety | 78 | 18.5% | 306 | 16.9% | 378 | 22.1% | 277 | 17.1% | 1,039 | 18.6% |
| Self-harm | 1 | 0.2% | 3 | 0.2% | 2 | 0.1% | 1 | 0.1% | 7 | 0.1% |
| Confusion | 24 | 5.7% | 141 | 7.8% | 133 | 7.8% | 119 | 7.3% | 417 | 7.5% |
| Depression | 126 | 29.9% | 452 | 24.9% | 396 | 23.2% | 407 | 25.1% | 1,381 | 24.8% |
| Mental state evaluation | 66 | 15.6% | 501 | 27.6% | 423 | 24.7% | 377 | 23.2% | 1,367 | 24.5% |
| Previous psychiatric history | 20 | 4.7% | 109 | 6.0% | 89 | 5.2% | 134 | 8.3% | 352 | 6.3% |
| Suicidal ideation | 2 | 0.5% | 8 | 0.4% | 9 | 0.5% | 3 | 0.2% | 22 | 0.4% |
| Insomnia | 8 | 1.9% | 51 | 2.8% | 85 | 5.0% | 98 | 6.0% | 242 | 4.3% |
| Suicide attempt | 0 | 0.0% | 1 | 0.1% | 4 | 0.2% | 2 | 0.1% | 7 | 0.1% |
| Behavioral problems | 9 | 2.1% | 27 | 1.5% | 12 | 0.7% | 16 | 1.0% | 64 | 1.1% |
| Psychosis | 8 | 1.9% | 9 | 0.5% | 22 | 1.3% | 19 | 1.2% | 58 | 1.0% |
| Treatment refusal | 3 | 0.7% | 5 | 0.3% | 6 | 0.4% | 20 | 1.2% | 34 | 0.6% |
| Psychopharmaceutical review | 33 | 7.8% | 73 | 4.0% | 52 | 3.0% | 45 | 2.8% | 203 | 3.6% |
| Somatization/Conversion | 1 | 0.2% | 9 | 0.5% | 13 | 0.8% | 16 | 1.0% | 39 | 0.7% |
| Follow-up | 3 | 0.7% | 13 | 0.7% | 23 | 1.3% | 10 | 0.6% | 49 | 0.9% |
| Discharge instructions | 3 | 0.7% | 11 | 0.6% | 6 | 0.4% | 1 | 0.1% | 21 | 0.4% |
| Recommendations of the Liaison Psychiatry team* |  |  |  |  |  |  |  |  |  |  |
| No requires attention | 55 | 13.0% | 145 | 8.0% | 186 | 10.9% | 263 | 16.2% | 649 | 11.6% |
| Initiation of medication | 270 | 64.0% | 1,123 | 61.8% | 1,033 | 60.4% | 1,047 | 64.5% | 3,473 | 62.3% |
| Change of medication dosage | 21 | 5.0% | 73 | 4.0% | 44 | 2.6% | 83 | 5.1% | 221 | 4.0% |
| Discontinue medication | 8 | 1.9% | 25 | 1.4% | 5 | 0.3% | 13 | 0.8% | 51 | 0.9% |
| Brief psychotherapy | 39 | 9.2% | 318 | 17.5% | 347 | 20.3% | 112 | 6.9% | 816 | 14.6% |
| Transfer to psychiatric hospitalization | 1 | 0.2% | 2 | 0.1% | 2 | 0.1% | 2 | 0.1% | 7 | 0.1% |
| Complete auxiliary exams | 7 | 1.7% | 34 | 1.9% | 18 | 1.1% | 7 | 0.4% | 66 | 1.2% |
| Requires the same treatment | 12 | 2.8% | 81 | 4.5% | 72 | 4.2% | 87 | 5.4% | 252 | 4.5% |
| Discharge from the liaison service | 9 | 2.1% | 15 | 0.8% | 3 | 0.2% | 10 | 0.6% | 37 | 0.7% |
| Psychiatric diagnoses** |  |  |  |  |  |  |  |  |  |  |
| F00-F09 Organic mental disorders including symptomatic disorders | 233 | 24.4% | 382 | 21.0% | 328 | 19.2% | 308 | 19.0% | 1,251 | 20.5% |
| F10-F19 Mental and behavioral disorders due to psychoactive substance use | 10 | 1.0% | 13 | 0.7% | 17 | 1.0% | 11 | 0.7% | 51 | 0.8% |
| F20-F29 Schizophrenia, schizotypal and delusional disorders | 38 | 4.0% | 39 | 2.1% | 26 | 1.5% | 31 | 1.9% | 134 | 2.2% |
| F30-F39 Mood (affective) disorders | 153 | 16.0% | 392 | 21.6% | 392 | 22.9% | 330 | 20.3% | 1,267 | 20.8% |
| F40-F48 Neurotic, stress-related and somatoform disorders | 458 | 48.0% | 759 | 41.8% | 680 | 39.8% | 699 | 43.0% | 2,596 | 42.5% |
| F50-F59 Behavioural syndromes associated with physiological disturbances and physical factors | 7 | 0.7% | 39 | 2.1% | 33 | 1.9% | 41 | 2.5% | 120 | 2.0% |
| F60-F69 Adult personality and behavioral disorders | 20 | 2.1% | 24 | 1.3% | 18 | 1.1% | 12 | 0.7% | 74 | 1.2% |
| F70-F79 Mental retardation | 5 | 0.5% | 3 | 0.2% | 4 | 0.2% | 6 | 0.4% | 18 | 0.3% |
| F80-F89 Developmental psychological disorders | 0 | 0.0% | 0 | 0.0% | 3 | 0.2% | 0 | 0.0% | 3 | 0.0% |
| F90-F98 Behavioural and emotional disorders often occur in childhood and adolescent | 0 | 0.0% | 1 | 0.1% | 2 | 0.1% | 0 | 0.0% | 3 | 0.0% |
| Z00-Z99 | 83 | 8.7% | 207 | 11.4% | 270 | 15.8% | 234 | 14.4% | 794 | 13.0% |
| Physical illnesses** |  |  |  |  |  |  |  |  |  |  |
| A00-B99 Certain infectious and parasitic diseases | 64 | 6.7% | 120 | 6.6% | 85 | 5.0% | 97 | 6.0% | 366 | 6.0% |
| C00-D48 Neoplasms | 191 | 20.0% | 364 | 20.0% | 344 | 20.1% | 363 | 22.4% | 1,262 | 20.7% |
| D50-D89 Diseases of the blood and hematopoietic organs and other disorders affect | 9 | 0.9% | 29 | 1.6% | 26 | 1.5% | 26 | 1.6% | 90 | 1.5% |
| E00-E90 Endocrine, nutritional and metabolic diseases | 78 | 8.2% | 152 | 8.4% | 165 | 9.6% | 158 | 9.7% | 553 | 9.1% |
| G00-G99 Diseases of the nervous system | 56 | 5.9% | 100 | 5.5% | 115 | 6.7% | 110 | 6.8% | 381 | 6.2% |
| H00-H59 Diseases of the eye and adnexa | 3 | 0.3% | 8 | 0.4% | 8 | 0.5% | 9 | 0.6% | 28 | 0.5% |
| H60-H95 Diseases of the ear and mastoid process | 2 | 0.2% | 3 | 0.2% | 5 | 0.3% | 3 | 0.2% | 13 | 0.2% |
| I00-I99 Diseases of the circulatory system | 86 | 9.0% | 168 | 9.3% | 138 | 8.1% | 120 | 7.4% | 512 | 8.4% |
| J00-J99 Diseases of the respiratory system | 62 | 6.5% | 227 | 12.5% | 172 | 10.1% | 112 | 6.9% | 573 | 9.4% |
| K00-K93 Diseases of the gastrointestinal tract | 44 | 4.6% | 112 | 6.2% | 144 | 8.4% | 139 | 8.6% | 439 | 7.2% |
| L00-L99 Diseases of the skin and subcutaneous tissues | 39 | 4.1% | 74 | 4.1% | 60 | 3.5% | 73 | 4.5% | 246 | 4.0% |
| M00-M99 Diseases of the musculoskeletal system and connective tissue | 39 | 4.1% | 88 | 4.8% | 100 | 5.8% | 78 | 4.8% | 305 | 5.0% |
| N00-N99 Diseases of the genitourinary system | 81 | 8.5% | 124 | 6.8% | 137 | 8.0% | 105 | 6.5% | 447 | 7.3% |
| O00-O99 Pregnancy, childbirth and puerperium | 14 | 1.5% | 9 | 0.5% | 12 | 0.7% | 23 | 1.4% | 58 | 1.0% |
| P00-P96 Certain conditions originating in the perinatal period | 0 | 0.0% | 0 | 0.0% | 1 | 0.1% | 1 | 0.1% | 2 | 0.0% |
| Q00-Q99 Congenital malformations, deformities and chromosome anomalies | 3 | 0.3% | 8 | 0.4% | 6 | 0.4% | 12 | 0.7% | 29 | 0.5% |
| R00-R99 Symptoms, signs, and abnormal clinical and laboratory findings, not else | 19 | 2.0% | 59 | 3.2% | 51 | 3.0% | 42 | 2.6% | 171 | 2.8% |
| S00-T98 Trauma, poisoning, and certain other consequences of external cause | 42 | 4.4% | 94 | 5.2% | 74 | 4.3% | 106 | 6.5% | 316 | 5.2% |
| V01-Y98 External causes of morbidity and mortality | 7 | 0.7% | 2 | 0.1% | 0 | 0.0% | 4 | 0.2% | 13 | 0.2% |
| Z00-Z99 Factors influencing health status and contact with healthcare services | 55 | 5.8% | 140 | 7.7% | 146 | 8.5% | 115 | 7.1% | 456 | 7.5% |
| U00-U99 Codes for special situations | 63 | 6.6% | 138 | 7.6% | 36 | 2.1% | 9 | 0.6% | 246 | 4.0% |
| Prescribed medication** |  |  |  |  |  |  |  |  |  |  |
| Antipsychotic | 278 | 29.1% | 555 | 30.6% | 394 | 23.0% | 413 | 25.4% | 1,640 | 26.9% |
| Antidepressant | 386 | 40.4% | 669 | 36.8% | 510 | 29.8% | 611 | 37.6% | 2,176 | 35.6% |
| Mood stabilizer | 32 | 3.4% | 51 | 2.8% | 45 | 2.6% | 58 | 3.6% | 186 | 3.0% |
| Anxiolytic | 433 | 45.3% | 764 | 42.1% | 696 | 40.7% | 655 | 40.3% | 2,548 | 41.7% |

Note: n=number, %=Percentage, \*Variables with missing data, \*\*Participants may have more than one diagnosis, so percentages may sum to more than 100%.

Table 2 presents the number of referrals by service. Notably, internal medicine recorded the highest number of referrals each year throughout the study period, with a peak of 546 referrals in 2024, representing 33.6% of all referrals to the liaison psychiatry service for that year. In contrast, services such as endocrinology and rheumatology had a lower number of referrals.

**Table 2.** Number of referrals per service and year (n=6,105).

| Referring Service | 2020<br>(n=955) |  | 2021<br>(n=1,816) |  | 2022<br>(n=1,710) |  | 2023<br>(n=1,624) |  | Total<br>(n=6,105) |  |
| --- | --- | --- | --- | --- | --- | --- | --- | --- | --- | --- |
|  | n | % | n | % | n | % | n | % | n | % |
| Medicine | 217 | 22.7% | 418 | 23.0% | 458 | 26.8% | 546 | 33.6% | 1639 | 26.8% |
| Pneumology | 66 | 6.9% | 224 | 12.3% | 179 | 10.5% | 96 | 5.9% | 565 | 9.3% |
| Others | 84 | 8.8% | 142 | 7.8% | 147 | 8.6% | 110 | 6.8% | 483 | 7.9% |
| General surgery | 50 | 5.2% | 132 | 7.3% | 94 | 5.5% | 128 | 7.9% | 404 | 6.6% |
| Pneumology | 63 | 6.6% | 125 | 6.9% | 108 | 6.3% | 101 | 6.2% | 397 | 6.5% |
| Geriatrics | 50 | 5.2% | 80 | 4.4% | 113 | 6.6% | 90 | 5.5% | 333 | 5.5% |
| Intensive Care Unit (ICU) | 21 | 2.2% | 110 | 6.1% | 67 | 3.9% | 45 | 2.8% | 243 | 4.0% |
| Gynecology | 65 | 6.8% | 58 | 3.2% | 50 | 2.9% | 66 | 4.1% | 239 | 3.9% |
| Gastroenterology | 24 | 2.5% | 70 | 3.9% | 53 | 3.1% | 66 | 4.1% | 213 | 3.5% |
| Traumatology | 29 | 3.0% | 63 | 3.5% | 29 | 1.7% | 59 | 3.6% | 180 | 2.9% |
| Dermatology | 28 | 2.9% | 42 | 2.3% | 58 | 3.4% | 43 | 2.6% | 171 | 2.8% |
| COVID | 58 | 6.1% | 88 | 4.8% | 19 | 1.1% | 0 | 0.0% | 165 | 2.7% |
| Cardiology | 28 | 2.9% | 45 | 2.5% | 45 | 2.6% | 41 | 2.5% | 159 | 2.6% |
| Neurosurgery | 27 | 2.8% | 35 | 1.9% | 45 | 2.6% | 45 | 2.8% | 152 | 2.5% |
| Nephrology | 33 | 3.5% | 33 | 1.8% | 38 | 2.2% | 45 | 2.8% | 149 | 2.4% |
| Renal transplant | 24 | 2.5% | 40 | 2.2% | 72 | 4.2% | 5 | 0.3% | 141 | 2.3% |
| Oncology | 27 | 2.8% | 39 | 2.1% | 40 | 2.3% | 27 | 1.7% | 133 | 2.2% |
| Obstetrics | 25 | 2.6% | 19 | 1.0% | 30 | 1.8% | 47 | 2.9% | 121 | 2.0% |
| Rheumatology | 23 | 2.4% | 32 | 1.8% | 31 | 1.8% | 27 | 1.7% | 113 | 1.9% |
| Endocrinology | 13 | 1.4% | 21 | 1.2% | 34 | 2.0% | 37 | 2.3% | 105 | 1.7% |

**Table 3.**
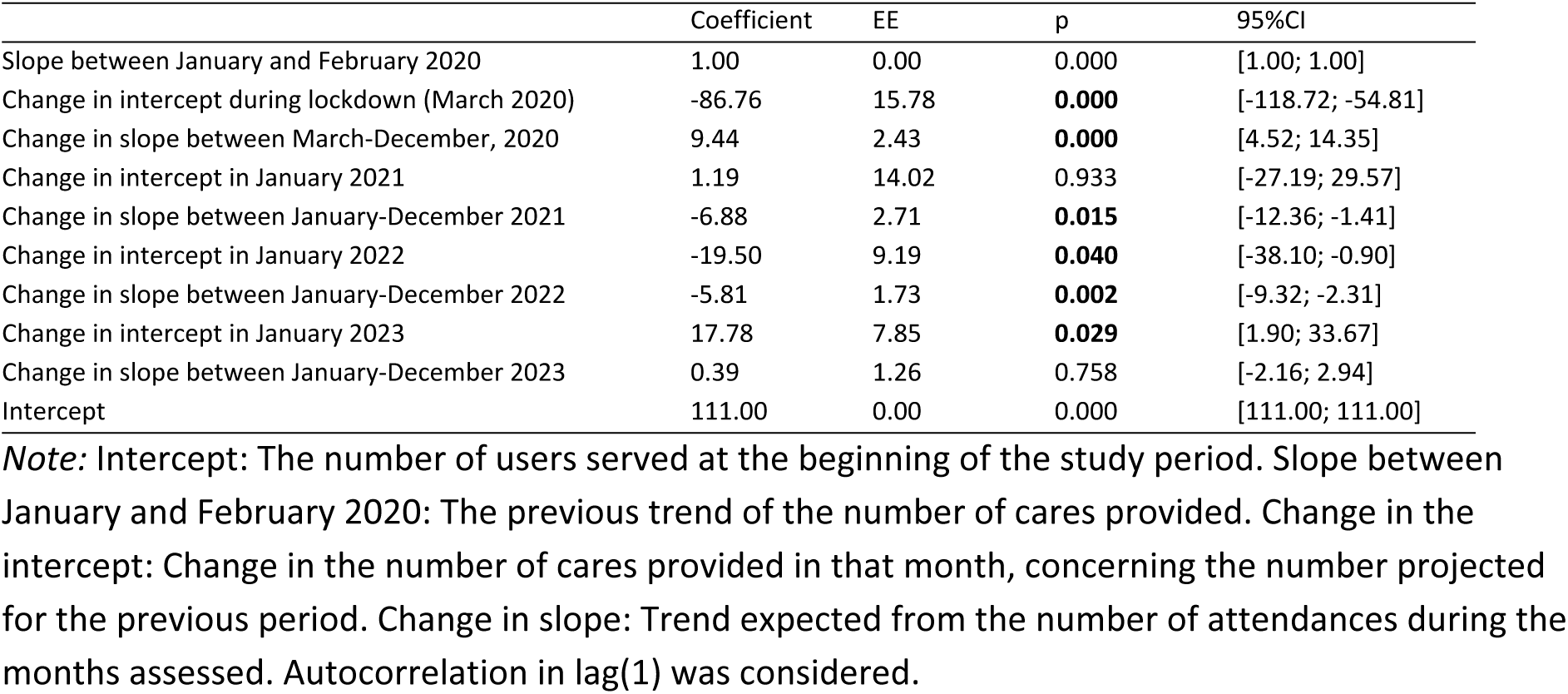
Interrupted time series analysis from 2020 to 2023 (n=6,105).

### Somatic and psychiatric diagnoses

The most common physical diseases among the referred cases were neoplasms (C00-D48) (n=14,262; 20.7%), diseases of the respiratory system (J00-J99) (n=573; 9.4%), and endocrine, nutritional, and metabolic diseases (E00-E90) (n=553; 9.1%). The most common psychiatric diagnoses were neurotic, stress-related and somatoform disorders (F40-F48) (n=2,596; 42.5%), affective disorders (F30-F39) (n=1,267; 20.8%), and organic mental disorders including symptomatic conditions (F00-F09) (n=1,251; 20.5%). The gender and year distributions for each diagnosis are shown in Supplementary material 3.

### Interrupted time series analysis

In March 2020, there was a decrease in the number of attendances compared to January and February of the same year (coeff=-86.76 [-118.72; –54.81]; p<0.001), a finding that could be attributed to the initial impact of the COVID-19 pandemic. During the remainder of 2020, there was a significant trend towards an increase in the number of visits per month (coeff=9.44 [4.52; 14.35]; p<0.001). However, in 2021, although there was an increase in the number of visits, a negative trend in such number per month was found (Figure 2), it was lower than expected when compared to previous months (coeff=-6.88 [-12.36; –1.41]; p=0.015). Similarly, in January 2022, a decrease in the number of care visits per month was observed (coeff=-19.50 [-38.10; –0.90]; p=0.040), compared to what was projected in the previous months, and during the year 2022, a decreasing monthly trend was found (coeff=-5.81 [-9.32; –2.31]; p=0.002). Finally, in January 2023, there was an increase in the number of care services provided when compared to the previous months (coeff=17.78 [1.90; 33.67]; p=0.029).

**Figure 2.** The number of cares provided between 2020 and 2023 (n=6,105).

## Discussion

### Main findings

To our knowledge, this is the first comprehensive analysis of psychiatry referral patterns to a Peruvian CLP Service over a four-year period, most probably conveying the profound disruptions caused by the COVID-19 pandemic. The overall referral rate of 6.73 % observed in the study is notably higher than those reported in other investigations. For instance, the finding in similar studies from Europe and Asia typically range between 1 to 5 % (6, 21, 22). Comparatively, studies from other countries report even lower referrals: 1.8 % in Italy (22), 2.2 % in Spain (21), 0.7 to 6 % in England (23), 1.19 % in Iran (24), 0.01 to 3.6 % in India (6), 0.02 to 3.6 % in China (25), and 3.3 % in Australia (26). The differences in healthcare systems, training, and in the integration of psychiatric services into general hospital settings might account for these variations. This discrepancy may be attributed to several factors unique to our setting at the HNGAI. The long-standing and prominent history of CLP at our institution, dating back to the late 1940s, has likely played a crucial role in sensitizing medical staff to the importance of psychiatric evaluations (14, 27). This historical context fosters a culture where psychiatric consultation is deeply integrated into the medical care process, and continuous academic and research activities in psychiatry, further enhance the positive disposition of internists, surgeons and other specialists to seek psychiatric consultation for their patients. Furthermore, all medical residents in Peru, regardless of their specialty, receive comprehensive mental health training, which facilitates recognition, appropriate attention, and discussion of psychiatric conditions, thereby contributing to more referrals observed (13). Additionally, this finding confirms the observations by Chen et al. regarding systemic factors that improve referral rates, such as a dedicated CLP service, active CLP consultants, and collaborative patient screening programs (28).

From the socio-demographic perspective, the high proportion of individuals over the age of 65 (40.1% of the referrals) highlights the crucial burden of mental health issues among older adults in general and within the realm of CLP. Certainly, the predominance of individuals over the age of 65 accounting for most hospital admissions is a plausible initial interpretation of these data. There are, however, additional considerations. This finding aligns with similar studies, such as one conducted in Canada, where 41.6 % of the patients were also found to be over the age of 65 (29), and another in Italy, where the proportion was 38.9 % (30). Additionally, the Italian study found that the number of referrals for older patients significantly increased over a 20-year period (30). This finding emphasizes the substantial need for focused attention and tailored interventions on older adults referred to CLPs: these patients frequently present complex clinical profiles characterized by multiple comorbidities, functional impairments, and psychosocial stressors (31). Mental health challenges include mood disorders, different modalities of cognitive impairment, substance use disorders, and adjustment difficulties related to chronic illness or bereavement (31, 32).

Interestingly, sex distribution within the sample shows some variability over the study period. While the proportion of men and women remained relatively balanced in 2020 and 2021, a notable decrease in the percentage of men was observed in 2022 and 2023, accounting then for only 39.9 % and 42.3 % of the sample, respectively. This shift raises questions about potential gender-specific factors influencing help-seeking behaviors or referral patterns within our healthcare system. Further investigation into societal norms, gender roles, and access to mental health resources may shed light on this phenomenon. There is considerable variability and lack of consensus regarding gender distribution within the field of CLP. Existing literature presents conflicting findings, with some studies reporting a predominance of referred male patients (5, 21, 33), while others show a higher proportion of females (22, 24). This inconsistency underscores the need for further research to elucidate the factors influencing gender differences in help-seeking behaviors and referral patterns within CLP contexts.

The number of patients referred to liaison psychiatry showed significant variability between 2020 and 2023 (see Figure 1), ranging from 111 patients (5.03% of total referrals) at the beginning of the study (January 2020) to a sharp decline of 15 patients (1.9% of total referrals) at the onset of the pandemic (April 2020). The number then progressively increased (n=132; 6.16% of total referrals), reaching a stable level by the end of the first year of the pandemic (January 2021). Subsequently, the number of referrals to liaison psychiatry remained constant. The initial sharp decline in early 2020 can be attributed to the onset of the COVID-19 pandemic, during which elective admissions and non-urgent consultations were postponed to prioritize acute care and resource allocation for COVID-19 patients. This phenomenon is consistent with findings from other studies documenting a reduction in healthcare utilization during the early pandemic period (34, 35). The subsequent increase and peak in referrals, reaching a maximum of 10.50 % in November 2021, likely correspond to the resumption of regular hospital activities and adaptation to pandemic-related challenges, such as enhanced infection control measures, and the implementation of telehealth services (35). Throughout 2021 and 2022, the referrals exhibited periodic fluctuations, potentially influenced by waves of COVID-19 infections and corresponding public health measures. For instance, spikes in referrals may align with periods of reduced COVID-19 cases, allowing for more consistent access to medical and psychiatric services. Conversely, declines may correspond with new waves of infections or the emergence of new variants, leading to renewed healthcare disruptions. The observed trends underscore the need for robust CLP services capable of adapting to fluctuating demands and external pressures.

The above considerations generate, in turn, questions such as the significant and well-known emotional/mental impact of COVID-19 on medical patients who require hospitalization for their general physical condition (36). Was the severity of COVID-19 cases such that it prevented physicians from requesting liaison psychiatry consultations? Or were the physicians from the other services simply overwhelmed by the care demands of an increased inpatient population? Research on these topics, even if retrospective, may show revealing features of hospital life in crisis situations.

### Public health implications

Understanding the profiles of inpatient referrals to the CLP Service of a general hospital has significant public health implications in countries like Peru and others with similar socio-economic contexts. By elucidating the patterns of psychiatric referrals and their interaction with medical conditions, this study provides relevant insights for healthcare policymakers and practitioners in resource-constrained settings.

First and foremost, a CLP clinical population provides a unique scenario for identification and management studies in the growing field of Social Determinants of Mental Health (37), not only for the mixed features of the patients but also for the convergence of objective and subjective factors, complications of etio-pathogenic mechanisms, and the dynamics triggered by personality, family, community and cultural ingredients (38).

Second, the findings underscore the importance of integrating mental health services within general hospital settings, especially those catering to a diverse range of medical and surgical conditions. This integration allows for the identification and management of psychiatric disorders among inpatients, leading to a better understanding of the clinical setting itself, and subsequently more comprehensive and holistic patient care (39).

Third, the study highlights the prevalence of certain psychiatric and somatic diagnoses, such as neurotic, stress-related, and somatoform disorders, as well as diseases of the respiratory system and neoplasms. Understanding these prevalent conditions can guide consistent healthcare resources allocation and an adequate use of available management routes, ensuring the most appropriate interventions and subsequent follow-up support.

Moreover, the analysis of referrals per service reveals variations across different medical specialties, indicating areas where targeted interventions may be needed to improve psychiatric care integration and referral practices. For instance, the consistently high referrals observed in Neurology and Geriatrics suggest a need for enhanced collaboration between psychiatric and such medical specialties to address the complex needs of patients with those types of comorbid conditions.

Furthermore, the interrupted time series analysis provides valuable insights into the impact of external factors, such as the COVID-19 pandemic, on psychiatric care provision and utilization. By identifying periods of change in the number of care visits, healthcare systems can adapt their policies and services to better respond to emerging challenges, such as the increased demand for mental health support during public health crises.

Finally, medical and public health education, training, and research activities in academic and training centers face both a challenge and an opportunity to improve didactic, instrumental, and evaluation approaches to CLP itself as well as psychiatry, medicine, and other health professions. By reasserting that the best clinical practice recognizes team and multidisciplinary work as its most solid component, CLP can become an ideal scenario for these pedagogical postulates (40). Research studies must go beyond descriptive or narrative approaches to explore truly bio-psycho-socio-cultural areas of CLP and its ontological routes and branches (41).

### Strengths and limitations

The study provides a comprehensive analysis of inpatient psychiatry referrals in a general hospital over a four-year period, shedding light on referral patterns and trends within a CLP Service. The utilization of rigorous statistical methods, such as interrupted time series analysis, enhances the robustness of the findings. Additionally, the study’s focus on a diverse range of sociodemographic and clinical characteristics of the probands enriches the understanding of this unique patient population, and opens the door for future, more complete research initiatives.

On the other hand, the retrospective design of reliance on existing medical records may create biases that could potentially result in incomplete or inaccurate data collection. Coming from a single-center study, the generalizability of the findings to other populations or settings may be limited. The utilization of a non-probabilistic sampling method and its eventual selection biases impact the representativeness of the sample and the applicability of the results. Moreover, the quality of data can be compromised by relying on the accuracy and completeness of medical documentation that may vary across healthcare centers and/or between departments of the same facility. In addition, although our study includes data on the number of referrals made to the liaison psychiatry service (numerator), it lacks information on the total proportion of patients referred to specific services or the specific flow of patients between different services (denominator).

### Conclusions

This study and analysis of the work of a Consultation Liaison Psychiatry Service within a large general hospital offers a unique opportunity to assess the clinical/emotional realities of medical and surgical inpatients and examine the dynamics of multiple interactions against an integrative background. Even a descriptive report of these activities shows unique clinical characteristics, socio-demographic aspects, and their relevance vis-à-vis diagnoses, management, and prognosis of both somatic and psychiatric conditions. Levels of integration of the CLP service staff with the referring services or departments, response, treatment measures, and their impact, follow-up, and outcomes of cases become well-defined research objectives. Innovative treatment modalities, i.e., psychotherapeutic approaches, and didactic/training strategies, particularly at post-graduate and specialization stages, are additional study foci and potential sources of excellence in the provision of CLP services in general hospitals.

## Declarations

### Availability of data and materials

The database can be requested from the corresponding author. The data comes from an electronic medical record, so it is not possible to share it due to privacy issues.

### Clinical trials registration numbers

Not applicable.

### Ethics approval and consent to participate

The Hospital Nacional Guillermo Almenara Irigoyen’s Institutional Review Board approved the protocol of this study (Carta N°122 CIEI-OIyD-GRPA-Essalud-2025). Throughout the process, the researchers had no access to identifying information about the participants. All participants were registered users of the hospital’s Liaison Psychiatry Service and received psychological or psychiatric care as needed. Participants did not need to provide informed consent, as the data was collected retrospectively. In addition, it was ensured that all data had been completely anonymized.

### Consent for publication

Not applicable.

## Acknowledgments

Not applicable.

## Declaration of generative AI and AI-assisted technologies in the writing process

We used DeepL to translate specific sections of the manuscript and Grammarly to improve the wording of certain sections. The final version of the manuscript was reviewed and approved by all authors.

## Authors’ contributions

**Jeff Huarcaya-Victoria:** Conceptualization, Methodology, Software, Validation, Formal analysis, Investigation, Resources, Data Curation, Writing – Review & Editing, Project administration.

**David Villarreal-Zegarra:** Conceptualization, Methodology, Software, Validation, Formal analysis, Data analysis, Writing – Review & Editing, Project administration.

**Renato D. Alarcón-Guzmán:** Conceptualization, Writing – Review & Editing, Project administration.

## Tables, Figures, and Supplementary Material

**Supplement material 1.** STROBE checklist.

**Supplement material 2.** Number and percentage of visits per month (n=6,105).

**Supplement material 3.** Percentage of somatic and psychiatric diagnoses: (A) and (B) and by year: (C) and (D) by sex.

